# “Unveiling Insulin Fear: Awareness, Myths and Practices Surrounding Injectable Insulin among working women suffering from Type 2 Diabetes Mellitus”

**DOI:** 10.1101/2024.07.09.24310042

**Authors:** Syed Rohan Ali, Aabir Imran, Aaila Haider, Kiran Mehtab, Muneeba Anees, Muhammad Hammad ul Haq, Muhammad Umer, Sawera Khan, Kainat Athar, Muhammad Hasan

**Affiliations:** Liaquat College of Medicine and Dentistry; Liaquat College of Medicine and Dentistry, Karachi; Jinnah Medical and Dental College, Karachi; Jinnah Sindh Medical University; Dow University of Health Sciences, Karachi

**Keywords:** Diabetes mellitus Type 2, Injections, Insulin, Discontinuation, Compliance, Fear, Psychological insulin resistance, primary care clinics, working women

## Abstract

**Objective:** This study aims to investigate the factors contributing to the reluctance in initiating and continuing insulin therapy among working women diagnosed with type 2 diabetes mellitus in karachi.

**Background:** A significant number of patients with type 2 diabetes do not achieve adequate control with maximum oral treatments. Early introduction of insulin has been shown to reduce diabetes-related complications. The purpose of this research is to assess the demographic characteristics, clinical factors, and attitudes of type 2 diabetes patients towards initiating insulin therapy. Currently, there is limited data available on insulin therapy for diabetes patients, particularly in Karachi. Therefore, additional randomized and prospective clinical trials are necessary to expand our knowledge and enable healthcare providers to make informed treatment decisions for diabetic patients in this region. Notably, there are various misconceptions surrounding insulin therapy, leading to refusal and delayed initiation, presenting a challenge to healthcare providers. Psychological insulin resistance is also prevalent among diabetic patients, hindering insulin therapy initiation and adherence.

**Methods:** A cross-sectional study will be conducted, involving a sample size of 167 diabetic female patients, determined using RAOSOFT software based on an estimated population size of 200,000. The sample will be selected through non-probability purposive sampling from Darul Sehat Hospital and healthcare clinics within an 8-month period from November 2023 to July 2024. Informed verbal consent will be obtained from each patient, and the validity of the questionnaire will be assessed. Data will be collected using a structured questionnaire distributed by the researcher. Statistical analysis will be performed using SPSS Version 22 with a 95% confidence interval, 5% margin of error, and a significance level set at 0.05. The relationship between insulin usage and demographic characteristics and clinical data will be evaluated using the χ2 or t-test and logistic regression, with age, educational status, religion, type of job, and diabetes control history as potential effect modifiers.

**Result:** The data highlights increasing insulin use with disease progression (p = 0.002) and its association with higher education (p = 0.003), indicating awareness. Challenges like injection discomfort among older patients (p = 0.004) and cultural beliefs affecting insulin use (p = 0.005) underscore the need for targeted education and support.

**Conclusion:** The findings of this study reveal that factors contributing to insulin noncompliance among patients with type 2 diabetes include illiteracy, non-diabetic treatment regimens, misconceptions, no private place for women, fear of cameras. Unhygienic environment. and irrational fear of insulin injections, medication costs, availability issues, concerns about long-term use, lack of family support, poor patient health, infrequent medical visits, and challenges related to communication and monitoring blood glucose levels.

## Introduction

Diabetes Mellitus Type 2 (T2DM) is a chronic metabolic disorder characterized by hyperglycaemia resulting from insulin resistance and/or inadequate insulin secretion. It is a global health concern, affecting millions of individuals worldwide and presenting a significant burden on healthcare systems. Among nations with a high prevalence of diabetes mellitus, Pakistan holds the 7th position. In 2007, Pakistan had around 6.9 million diabetes mellitus patients, a number anticipated to rise to 11.5 million by 2025 according to the International Diabetic Foundation.^1^ Because type-2 Diabetes Mellitus is a condition that advances over time, mismanagement of oral-hypoglycemic medications result in initiation of insulin therapy act as a resort management option to attain the required glycemic targets and avert complications. Despite the well-established success of starting insulin treatment, patients frequently manifest significant psychological, cultural and religious reluctance, thereby adding complexity to the procedure.^4^ However, despite its proven efficacy, insulin utilization remains sub-optimal in many T2DM patients.^6^ A range of factors contributes to this inadequate utilization, spanning patient-related, cultural believes related, and system-related barriers.

The theory suggests that individual beliefs about insulin play a substantial role in determining the willingness to adopt and follow insulin therapy.^7^ Pakistani women face distinct challenges related to insulin injections, particularly those who are part of the workforce and struggle with concerns related to privacy and assistance. The phobia surrounding injections and unfavorable viewpoints, like the fear of harm to organs, impede acceptance. Work timings and environment also acts as a barrier in utilization of insulin among working women. The fear of being watched, issues with privacy, the unhygienic condition of the place also plays a role in people not opting insulin. Religious barriers are also a big factor as injection insulin is considered self harm^8^. Dress codes for different places and religious dressings also causes hindrance in injecting insulin in a public places. The maintenance and availability of insulin to a women in Pakistan may also be a factor due to which women opt for oral drugs.^9^ Likewise, anxieties about potential side effects and the possibility of feeling embarrassed hinder the commitment to adherence.

These convictions are molded by cultural and societal surroundings, molding themselves based on personal encounters. Nevertheless, there has been a noticeable absence of a recent study at the local level that delves into this aspect. Therefore, the present research strives to investigate the knowledge and myth regarding insulin phobia in patients of type 2 DM among working women of Karachi.

### Methodology

This descriptive cross-sectional study will be conducted over approximately eight months following Ethical approval from LCMD, in accordance with the Helsinki Declaration guidelines, focusing on the Type 2 diabetic working female population. A sample size of 167 diabetic female patients, derived using RAOSOFT software based on an estimated population size of 200,000, will be selected through non-probability purposive sampling. The questionnaire used for data collection has a Cronbach’s alpha value of 0.745. Data will be gathered by the researcher using a structured questionnaire, with an equal distribution of questionnaires among researchers and adherence to standardized operating procedures to minimize assessment bias.Statistical analysis will be performed using IBM SPSS Statistics Version 22, with a 95% confidence interval, a 5% margin of error, and a significance level of 0.05. The relationship between insulin usage and demographic characteristics and clinical data will be assessed using the χ2 or t-test and logistic regression, considering age, educational status, religion, type of job, and diabetes control history as potential effect modifiers.

## Result

The analysis of diabetic medication usage across various demographic categories reveals significant associations. Across different age groups, substantial variations were observed in medication preferences (p = 0.000). In the 30-40 age bracket, oral medication was favored by 46 patients, while 16 opted for insulin and 1 used both. Conversely, the 41-50 age group showed a preference for insulin with 37 patients, compared to 21 using oral medication and 2 using both. The 50-60 age group demonstrated a more balanced usage, with 17 patients on oral medication, 19 on insulin, and 8 using both forms. Religious affiliations also showed notable distinctions in medication choices (p = 0.009), with Muslims predominantly using oral medication (77 patients) and insulin (67 patients), whereas Hindus and Christians exhibited lower usage overall. Educational status significantly influenced medication patterns (p = 0.001), with higher educational attainment correlating with more insulin usage and lower reliance on oral medication. Job type (p = 0.000) and the duration of diabetes diagnosis (p = 0.002) further highlighted varying medication preferences, reflecting broader socio-economic and health management factors among diabetic patients.

The data shows significant insights into diabetic patients’ perceptions and fears regarding insulin usage across various demographics, supported by statistically significant findings (p < 0.05). Over time, there’s an increase in insulin usage with disease progression (p = 0.002). Higher education correlates with more insulin use (p = 0.003), reflecting awareness of treatment options. Older patients find insulin self-injection more challenging (p = 0.004). Educational attainment affects beliefs about cultural forbiddance of insulin use (p = 0.005). Married individuals report more discomfort with insulin injections in traditional attire (p = 0.006). Job type impacts perceptions of logistical barriers (p = 0.018). Younger patients fear weight gain from insulin use (p = 0.001). Beliefs about insulin’s long-term complications vary by age and duration of diabetes (p = 0.002), suggesting a need for tailored education and support in diabetes management.

## Discussion

The study provides a comprehensive analysis of diabetic medication usage among working female Type 2 diabetic patients, highlighting significant patterns across various demographic variables. The data reveals notable insights into the distribution and perceptions of oral medication and insulin use, influenced by age, religion, educational status, job type, and the duration of diabetes diagnosis.The analysis shows a significant variation in medication usage among different age groups (p-value = 0.000). In the 30 to 40 age group, a majority (46 out of 63) rely on oral medication, with fewer patients using insulin (16) or both (1). This trend shifts in the 41 to 50 age group, where insulin usage surpasses oral medication (37 vs. 21), and a similar pattern is observed in the 50 to 60 age group. This shift towards insulin with increasing age could be attributed to the progression of diabetes and the reduced efficacy of oral medications over time. The total distribution shows that out of 167 patients, 84 use oral medication, 72 use insulin, and 11 use both.

The study indicates a statistically significant difference in medication usage among different religious groups (p-value = 0.009). Among the 152 Muslim patients, oral medication is the most common (77), followed by insulin (67), and a few use both (8). In contrast, Hindu and Christian patients show a similar preference for oral medication but in smaller numbers due to the lower population size. This variation might reflect cultural or socioeconomic factors influencing treatment preferences and access to healthcare resources.Educational status also significantly affects medication usage (p-value = 0.001). Illiterate patients predominantly use oral medication (10 out of 14), while those with primary education show a higher propensity for insulin (27 out of 41). Secondary education level patients display a balanced usage between oral medication and insulin, whereas those with education above secondary level primarily use oral medication (44 out of 70). This trend suggests that higher educational status may correlate with better disease management knowledge and access to a variety of treatment options.

Job type significantly influences medication usage patterns (p-value = 0.000). Office workers exhibit a balanced use of oral medication and insulin, while maids show a higher reliance on oral medication (23 out of 35). Teachers tend to prefer oral medication (30 out of 45), and those in the “others” category primarily use insulin (33 out of 43). These variations may be influenced by job-related stress, physical activity levels, and access to healthcare facilities.The duration of diabetes diagnosis significantly affects medication usage (p-value = 0.002). Patients diagnosed within the last 1-5 years predominantly use oral medication (50 out of 90), while those diagnosed for 5-10 years show an equal preference for oral medication and insulin.Patients with a longer duration of diabetes (10-20 years) exhibit a higher usage of both insulin and oral medication, suggesting a need for more intensive management as the disease progresses.

Perceptions of insulin usage reveal significant trends across various demographic factors. Educational status significantly affects insulin usage (p-value = 0.003), with higher educational levels correlating with greater insulin use. Age also influences the perception of insulin self-injection difficulty (p-value = 0.004), with the 50-60 age group finding it more challenging compared to younger patients. Similarly, the duration of diabetes diagnosis affects perceptions of self-injection difficulty (p-value = 0.000), with newer patients (1-5 years) finding it harder than those diagnosed for longer periods. Cultural beliefs significantly impact insulin usage (p-value = 0.005). Illiterate and primary-educated patients are more likely to believe that insulin is forbidden in their culture compared to those with higher education. Marital status also influences perceptions of discomfort when injecting insulin while wearing an abaya (p-value = 0.006), with married individuals reporting higher discomfort levels. These findings highlight the need for culturally sensitive education and support to address misconceptions and improve insulin adherence.

Fear of long-term complications from insulin usage is significantly associated with age (p-value = 0.002) and duration of diabetes diagnosis (p-value = 0.000). Younger patients (30-40) and those newly diagnosed (1-5 years) are more likely to believe that insulin causes long-term complications, reflecting a need for better education on insulin’s safety and efficacy.Job type significantly affects perceptions of the feasibility of continuing insulin usage (p-value = 0.018). Office workers, maids, and teachers report challenges in continuing insulin treatment due to work timings, highlighting the need for flexible treatment plans and workplace support to ensure adherence.

## Conclusion

In conclusion, this study underscores the complex interplay between demographic factors and diabetic medication usage among working female patients. The significant variations observed across age, religion, educational status, job type, and duration of diabetes diagnosis emphasize the need for tailored interventions to address specific barriers and improve diabetes management outcomes. Comprehensive education, culturally sensitive support, and flexible treatment plans are crucial in enhancing medication adherence and overall health among diabetic patients.

## Data Availability

Which will made available on request

## Declarations

### Author Contributions

- **Concept & Design of Study:** Syed Rohan Ali
- **Drafting:** Aabir Imran
- **Data Analysis:** Syed Rohan Ali
- **Revisiting Critically:** Syed Rohan Ali
- **Final Approval of Version:** Aaila Haider, Muneeba Anees, and Kiran Mehtab

Syed Rohan Ali, Aabir Imran, Aaila Haider, Kiran Mehtab, Muneeba Anees, Muhammad Hammad ul Haq, Muhammad Umer, Sawera Khan, Kainat Athar, Muhammad Hasan played crucial roles in refining the content, conducting a final round of editing, and presenting the article in a structured and balanced format, ensuring it was ready for publication.

### IRB Reference No

IRB-66/04/ LCMD/08/2022

### Ethics Approval

Ethics approval was granted by the IRB Committee LCMD, Karachi.

### Consent

Verbal consent was obtained from the participants.

### Data Availability

Data was collected by the investigator. If required, data and materials access can be granted.

